# Where is Omicron? Comparison of SARS-CoV-2 RT-PCR and Antigen Test Sensitivity at Commonly Sampled Anatomic Sites Over the Course of Disease

**DOI:** 10.1101/2022.02.08.22270685

**Authors:** Jessica Lin, Jennifer K Frediani, Gregory L Damhorst, Julie A Sullivan, Adrianna Westbrook, Kaleb McLendon, Tyler J Baugh, William H O’Sick, John D Roback, Anne L Piantadosi, Jesse J Waggoner, Leda Bassit, Anuradha Rao, Morgan Greenleaf, Jared W O’Neal, Seegar Swanson, Nira R Pollock, Greg S Martin, Wilbur A Lam, Joshua M Levy

## Abstract

**Background:** Upper respiratory samples for SARS-CoV-2 detection include the gold standard nasopharyngeal (NP) swab, and mid-turbinate (MT) nasal swabs, oropharyngeal (OP) swabs, and saliva. Following the emergence of the omicron (B.1.1.529) variant, limited preliminary data suggest that OP swabs or saliva samples may be more sensitive than nasal swabs, highlighting the need to understand differences in viral load across different sites.

**Methods:** MT, OP, and saliva samples were collected from symptomatic individuals presenting for evaluation in Atlanta, GA, in January 2022. Longitudinal samples were collected from a family cohort following COVID-19 exposure to describe detection of viral targets over the course of infection.

**Results:** SARS-CoV-2 RNA and nucleocapsid antigen measurements demonstrated a nares-predominant phenotype in a familial cohort. A consistent dominant location for SARS-CoV-2 was not found among 54 individuals. Positive percent agreement for virus detection in MT, OP and saliva specimens were 66.7 [54.1–79.2], 82.2 [71.1–93.4], and 72.5 [60.3–84.8] by RT-PCR, respectively, and 46.2 [32.6–59.7], 51.2 [36.2–66.1], and 72.0 [59.6–84.4] by ultrasensitive antigen assay. The composite of positive MT or OP assay was not significantly different than either alone for both RT-PCR and antigen assay (PPA 86.7 [76.7–96.6] and 59.5 [44.7–74.4], respectively).

**Conclusions:** Our data suggest that SARS-CoV-2 nucleocapsid and RNA exhibited similar kinetics and diagnostic yield in three upper respiratory sample types across the duration of symptomatic disease. Collection of OP or combined nasal and OP samples does not appear to increase sensitivity versus validated nasal sampling for rapid detection of viral antigen.

## Introduction

Nasopharyngeal (NP) sampling by a trained healthcare professional is the gold standard for the diagnosis of SARS-CoV-2 infection.^1^ Due to the complexity of NP sampling and associated discomfort, anterior nares (AN), nasal mid-turbinate (MT), and oropharyngeal (OP) swabs and expectorated saliva have emerged as commonly employed sample types for some assays.^2–8^ AN and MT sampling are employed in at-home rapid antigen tests with US Food and Drug Administration emergency use authorization.^9^ The emergence of the omicron (B.1.1.529) variant, regional differences in sampling practices, and recent social media trends have reignited debate over the most effective method for sample collection in persons suspected of having COVID-19.^10,11^

Early meta-analyses of data corresponding to the alpha (B.1.1.7) variant demonstrated the highest sensitivity in NP swabs or combined nasal-OP swabs.^4,12–14^ Following the emergence of delta (B.1.617.2) and omicron (B.1.1.529) variants, there are minimal data addressing the relative clinical sensitivity of sample types. Meanwhile, there are few systematic comparisons of the impact of sample type on rapid antigen test sensitivity, limiting understanding of optimal sampling techniques.^15,16^

The omicron variant has over 50 mutations relative to the wild-type virus, which has been postulated to affect tropism and the anatomic distribution of virus in the upper respiratory tract, and the prevalence of certain symptoms such as odynophagia.^17^ Recent evidence suggests omicron replicates more easily in the bronchial compared to lung parenchymal tissues, which contrasts with measurements of delta and wild-type viruses.^18^ This highlights differences in tissue-dependent replication profiles that may be present in the upper airways, thus impacting diagnostic test performance. One preprint study shows that the omicron variant may be more readily detected in saliva compared to MT samples by RT-PCR,^19^ while another showed lower sensitivity of OP compared to AN swab.^20^

To inform diagnostic sampling practices in both home and point of care settings, we sought to quantify antigen and molecular test performance across three sample types among patients infected with SARS-CoV-2 during the recent omicron surge. Findings from a familial cohort were first examined in a longitudinal qualitative analysis. A cross-sectional cohort was analyzed to determine the diagnostic performance of samples collected from MT, OP, and saliva, as defined by differences in threshold cycle (Ct) value and quantitative antigen measurements. Additionally, we seek to evaluate the impact of symptom duration and vaccination status on test performance at each site.

## Methods

This study was approved by the Emory University Institutional Review Board under STUDY00001082. Recruitment was completed during the height of the omicron surge in the metro Atlanta, GA area in January 2022.

### Longitudinal sampling of a familial cohort

Following exposure of two related individuals to a known case of COVID-19, both individuals and a third family member were verbally consented over the phone and provided with self-collection kits for AN, OP, and saliva samples. Each participant provided almost daily specimens, which were refrigerated immediately and transported on ice to the Emory/Children’s Laboratory for Innovative Assay Development (ELIAD) for analysis within 72 hours of collection. AN and OP swabs were immediately placed in 3 mL saline, while saliva was collected using the SalivaDirect unsupervised collection kit (New Haven, CT), including a short straw (Salimetrics Saliva Collection Aid (Carlsbad, CA) and a sterile 2 mL plastic tube containing 3 ceramic beads.

### Testing site sample collection

MT, OP, and passive expectorated saliva samples were collected consecutively from individuals at community and hospital-based sites affiliated with Emory University Hospital, Grady Memorial Hospital, and Children’s Healthcare of Atlanta as part of the RADx program.^21^. Participants were included if they experienced COVID-19 symptom onset within the prior 7 days and consented to the collection of MT, OP and passive saliva specimens. Exclusion criteria included asymptomatic patients, those with symptoms associated with COVID-19 for greater than 7 days, and those unable to provide informed consent. MT and OP specimens were obtained by a healthcare provider trained with CDC guidance materials using a specimen swab (Mantacc, Miraclean Technology Co., Ltd. Shenzhen, China) and placed in 3 mL saline. Saliva specimens were collected using the SalivaDirect unsupervised collection kit (New Haven, CT) under the supervision of a healthcare provider, including a short straw (Salimetrics Saliva Collection Aid (Carlsbad, CA), catalog #5016.02) and a sterile 2 mL plastic tube containing 3 ceramic beads. All specimens were transported on ice to ELIAD for analysis within 24 hours of sample collection. Clinical and demographic variables were collected in a centralized, web-based database (REDcap, Nashville, TN).

### RT-PCR Testing

RT-PCR testing for MT and OP samples was performed using the Cepheid GeneXpert Dx Instrument System with Xpert Xpress CoV-2/Flu/RSV plus cartridges (EUA 302-6991, Rev. B., October 2021). Only results from SARS-CoV-2 are included herein. The reported PCR cycle threshold (Ct) value represents the first target to amplify from one of the three unique SARS-CoV-2 genome target sequences: nucleocapsid (N), envelope (E), and RNA-dependent RNA polymerase (RdRP). Assay result and Ct value were retrieved from GeneXpert System Software.

Saliva samples were homogenized at 4.5 m/s for 5 seconds using the Omni International Bead Ruptor Elite (Kennesaw, GA). In accordance with the SalivaDirect dual-plexed RT-qPCR protocol, 2.5 µL proteinase K was added to 50 µL homogenized saliva. Following proteinase K inactivation, 5 µL of the saliva-proteinase K preparation was added to reaction mix for a total reaction volume of 20 µL. The reaction was performed on the QuantStudio platform (ThermoFisher Scientific, Waltham, MA) ^22–24^, using reagents and materials qualified for the SalivaDirect procedure.

### Antigen Concentration

Nucleocapsid antigen was quantified using the SARS-CoV-2 N Protein Advantage assay on the Quanterix Simoa HD-X platform (Billerica, MA). Specimen residuals were stored at 4°C during Quanterix antigen screening. Samples were resulted positive for an antigen (cutoff >/=3.00pg/mL).

### Lateral Flow Assay Testing

In a subset of participants, an additional OP swab (collected simultaneously with the swab for RT-PCR and nucleocapsid testing) and one AN swab were collected by a healthcare provider for use with the Quickvue OTC home test (Quidel, San Diego, CA). The test was performed according to the manufacturer’s instructions by study staff within four minutes of sample collection.

### Statistical Analysis

Antigen concentrations and Ct values from MT, OP, and saliva sample testing were compared for 54 participants who tested positive for SARS-CoV-2 RNA or nucleocapsid protein in at least one of the sample types. Not all participants were able to contribute all three sample types. For the mixed effects model, samples that resulted negative for antigen were assigned a value of 2.99 pg/mL (immediately below the assay cut-off) and negative RT-PCR results were assigned a Ct value of 50 (above the highest Ct value observed in the data).

Absolute agreement of Ct values and antigen concentrations across sample types were calculated via an intraclass correlation coefficient (ICC) using a two-way mixed effects model in IBM SPSS Statistics for Windows, version 28 (IBM Corp., Armonk, N.Y., USA).

To examine the association between location and duration of symptoms on mean Ct value and antigen concentrations, Ct value and antigen concentrations were log transformed for normality and then analyzed in a mixed effects model accounting for correlation between measures from the same subjects in our cross-sectional cohort. Subjects were treated as a random effect while time and location were considered fixed effects. A model assuming independence and models assuming correlation between observations were applied to each outcome and a likelihood ratio test between nested models was employed to determine the model of best fit which would be used for the final analysis. Three possible correlation structures were considered: autoregression, Toeplitz, and unstructured correlation. The highest-level correlation structure that returned a significantly better fit than the previous structure was used for analysis. The adjusted back-transformed means of main effects, respective 95% confidence intervals, and F-test p-values were reported. This analysis was conducted in SAS 9.4 (Cary, NC).

## Results

### Familial cohort

In longitudinal self-collection of a familial cohort (n=3), each of the participants provided 8-10 samples over a period of 8-11 days beginning with the first day after symptom onset (Figure 1A-C). Two participants provided the results of daily point-of-care (POC) diagnostics tests. Overall, we observed consistently higher antigen concentrations and lower Ct values in AN samples compared to paired OP and saliva samples at all timepoints (nares-predominant phenotype; **Figure 1**). Relative to symptom onset, the first positive POC test was seen on different days for each participant (one day prior, one on the same day and one day following symptom onset).

**Figure 1.**
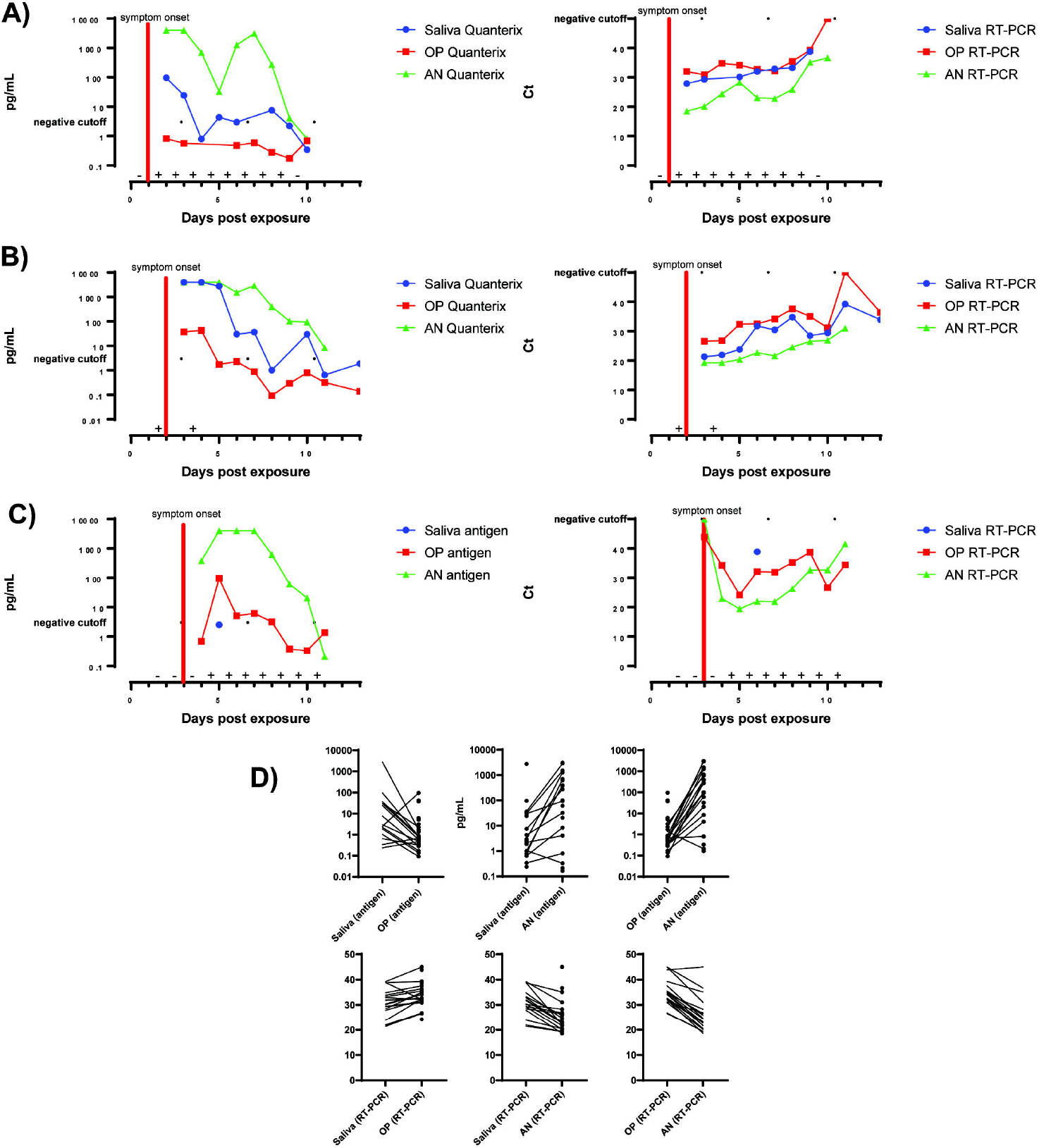
Comparison of antigen concentration and Ct value of different sample types over the course of disease in a familial cohort. A-C) Three COVID-19 positive patients from the same family cohort were tracked from the time of exposure over the course of disease. Each family member (A, B, or C) self-collected AN, OP, and saliva samples, which are measured using RT-PCR and the Quanterix Simoa HD-X. The time of symptom onset is indicated by a red line for each patient. Results of self-administered lateral flow assays are indicated as positive (+) or negative (-) on the x-axis. All negative RT-PCR tests are assigned a Ct value of 50 (indicated by the dotted line), which is above the highest detected Ct. The antigen concentration measurements too high to be quantified are assigned a value of 4000pg/mL, which is above the highest detected antigen level. D) Matched samples are paired according to sample type and the differences in antigen concentration (top) and Ct value (bottom) are shown.

### Cross-sectional cohort participants and characteristics

MT, OP, and saliva specimens were collected from 121 individuals in the cross-sectional cohort of whom 54 tested positive for SARS-CoV-2 by RT-PCR from one or more specimens. 29 (53.7%) of the cohort was female and 45 (83.3%) had received at least one dose of a COVID-19 vaccine (**Table 1**). Median days since symptom onset was 2.0 (IQR 1.0 – 3.0). The most common symptoms at the time of evaluation were congestion or rhinorrhea (31, 57.4%) and cough (30, 55.6%; **Table 1**). Odynophagia was reported by 20 (37.0%) of patients at time of evaluation. Only one participant reported having ageusia or anosmia.

**Table 1.**
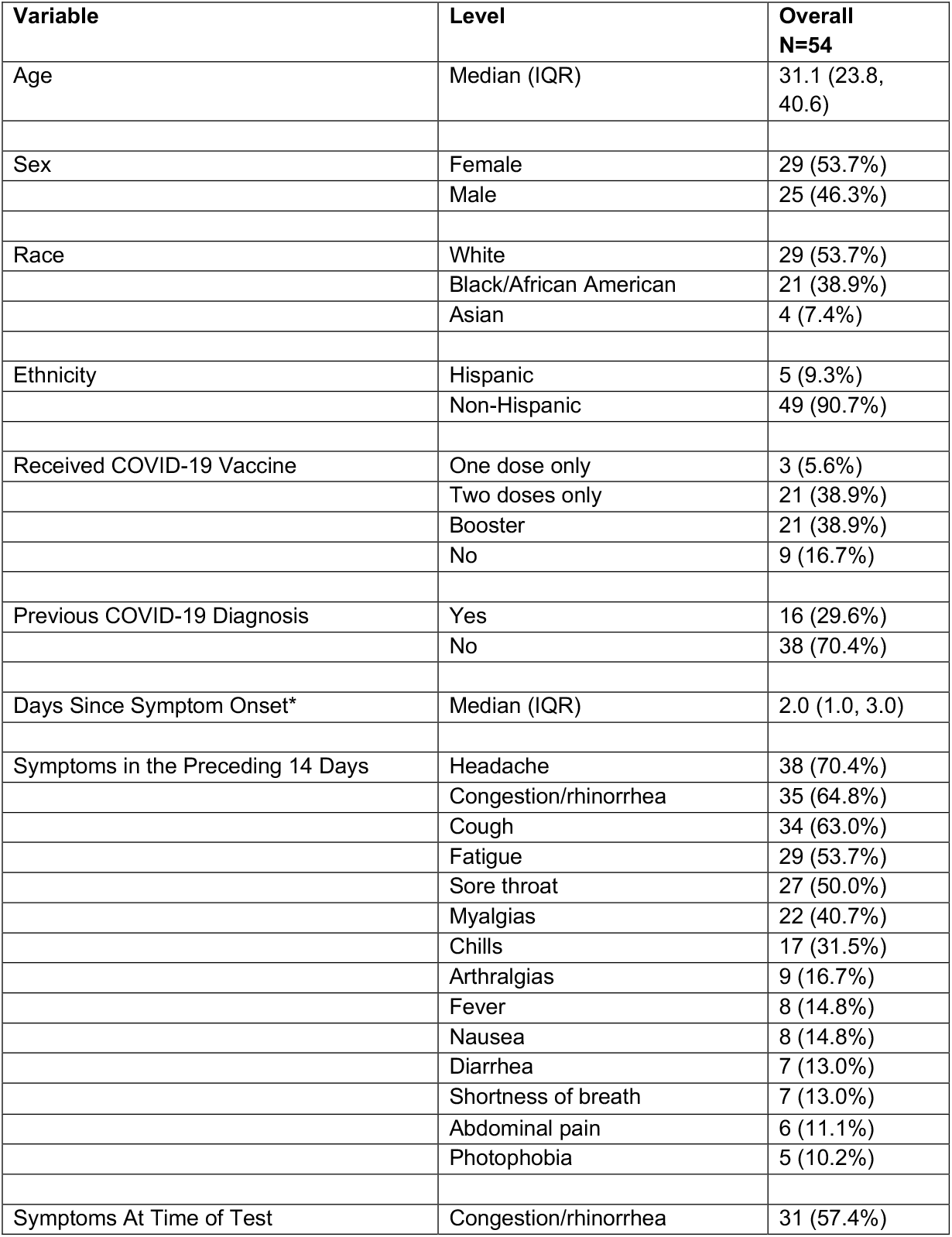
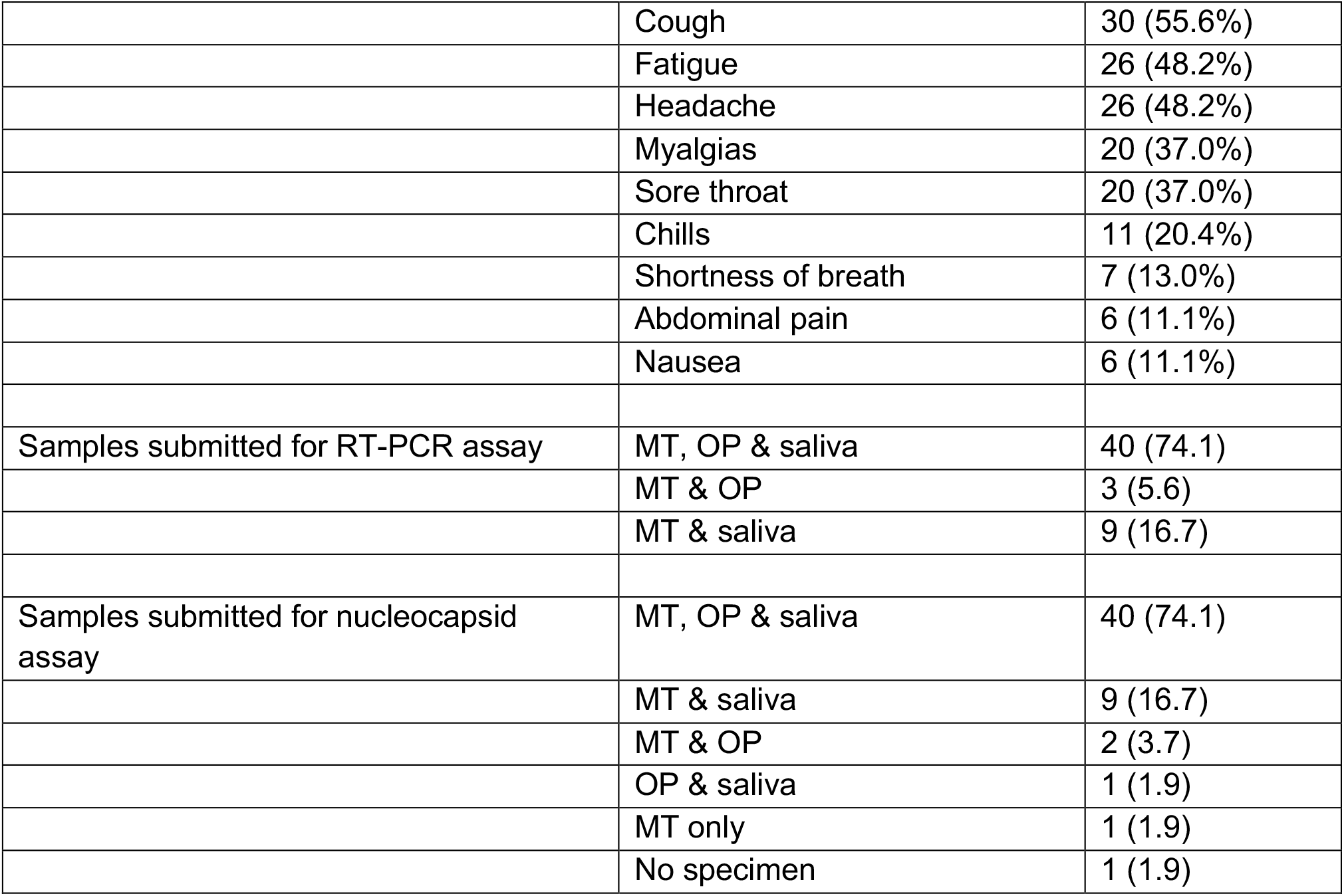
Characteristics of study population and sample collection. Symptoms with less than 10% prevalence are omitted. *Those who reported no symptoms were excluded from this calculation

### Diagnostic performance

RT-PCR was performed on 54 MT, 45 OP, and 51 saliva specimens while nucleocapsid antigen assay was performed on 52 MT, 43 OP and 50 saliva specimens (**Table 1**). PPA [95% CI] for RT-PCR was 66.7 [54.1–79.2], 82.2 [71.1–93.4], and 72.5 [60.3–84.8] in MT, OP and saliva specimens, respectively, while antigen detection exhibited 46.2 [32.6–59.7], 51.2 [36.2–66.1], and 72.0 [59.6–84.4] PPA at a cutoff of 3 pg/mL (**Figure 2A**). A composite result of MT or OP specimens, which was calculated only for individuals who had both MT and OP specimens analyzed, showed 86.7 [76.7–96.6] for RT-PCR and 59.5 [44.7–74.4] for antigen detection.

**Figure 2.**
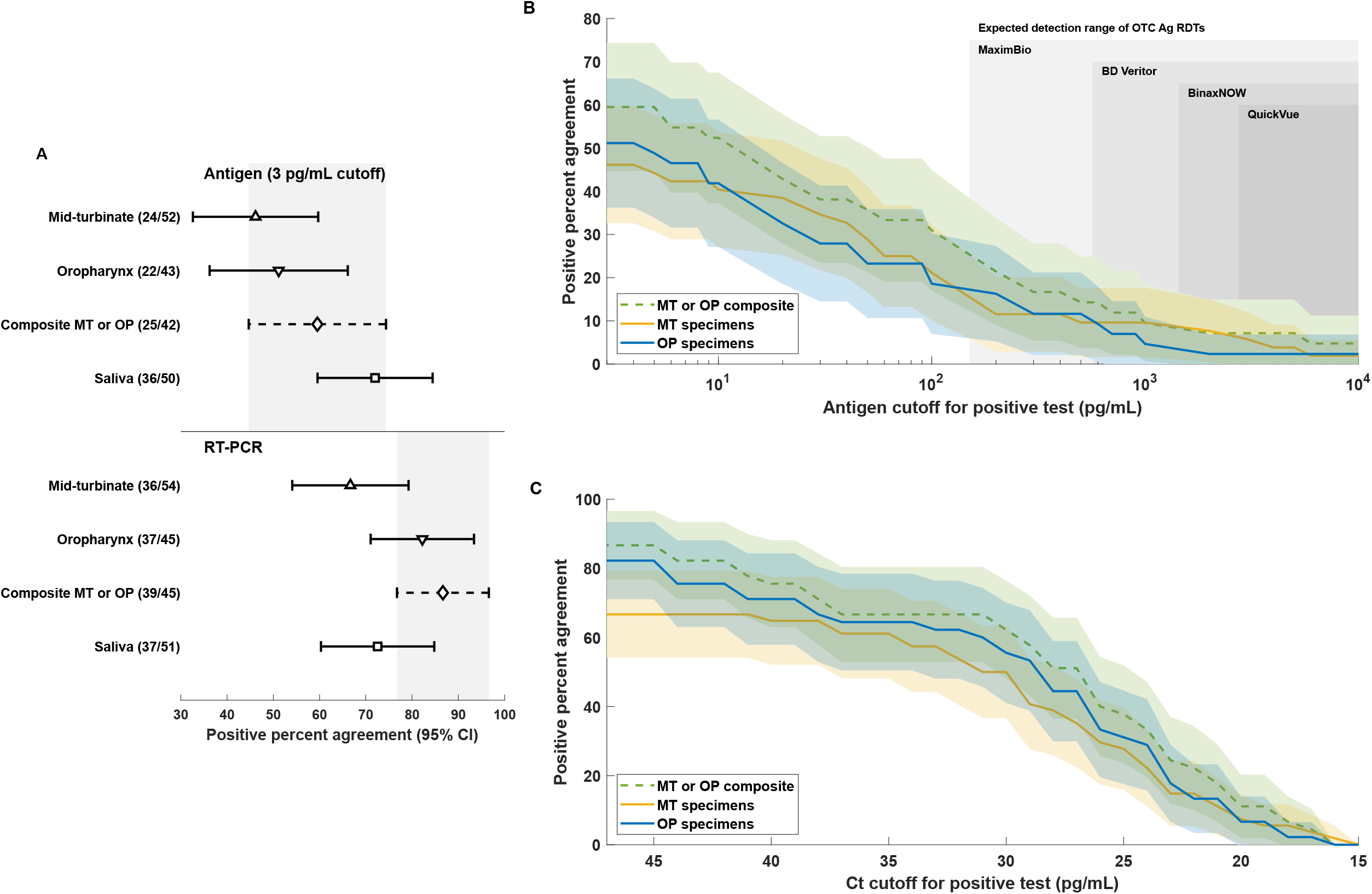
Positive percent agreement (PPA) of RT-PCR and antigen measurements of different specimen types. All included specimens came from participants who had one or more of the three specimens positive for SARS-CoV-2 by RT-PCR. Results are included in the composite MT or OP only if the participant had both samples available. (A) Antigen PPA determined by a cutoff of 3 pg/mL and RT-PCR (threshold cycle cutoff based on respective assay). (B) Antigen PPA for MT, OP and MT or OP composite for a range of antigen cutoffs. Shaded regions denote 95% CI calculated using the formula for standard error. Expected ranges for commercially available EUA rapid diagnostic tests are shown in gray (see Table S1). (C) RT-PCR PPA for MT, OP and MT or OP composite versus threshold cycle cutoff. Shaded regions denote 95% CI calculated using the formula for standard error.

In examination of the diagnostic performance of antigen detection at varying cut-offs, significant overlap was observed within the 95% CI for PPA of MT, OP sample types, and the composite result of MT or OP. These calculations are plotted over reference ranges of common over the counter (OTC) rapid diagnostic tests (RDTs) for reference in **Figure 2B**. Similarly, PPA for RT-PCR over the range of Ct values is shown in **Figure 2C**.

### Viral load distribution and agreement between sample types

Distribution of Ct value, a surrogate marker for viral load, did not show significant differences in comparison of any two sample types across the cohort (MT vs OP p = 0.32, SA vs OP p = 0.26, MT vs SA, p = 0.8; **Figure 3A**). Nucleocapsid antigen measurements were significantly different in comparison of saliva with MT or OP samples (p = 0.02 and 0.03, respectively; **Figure 3B**), but not in the comparison of MT and OP samples (p = 0.88).

**Figure 3.**
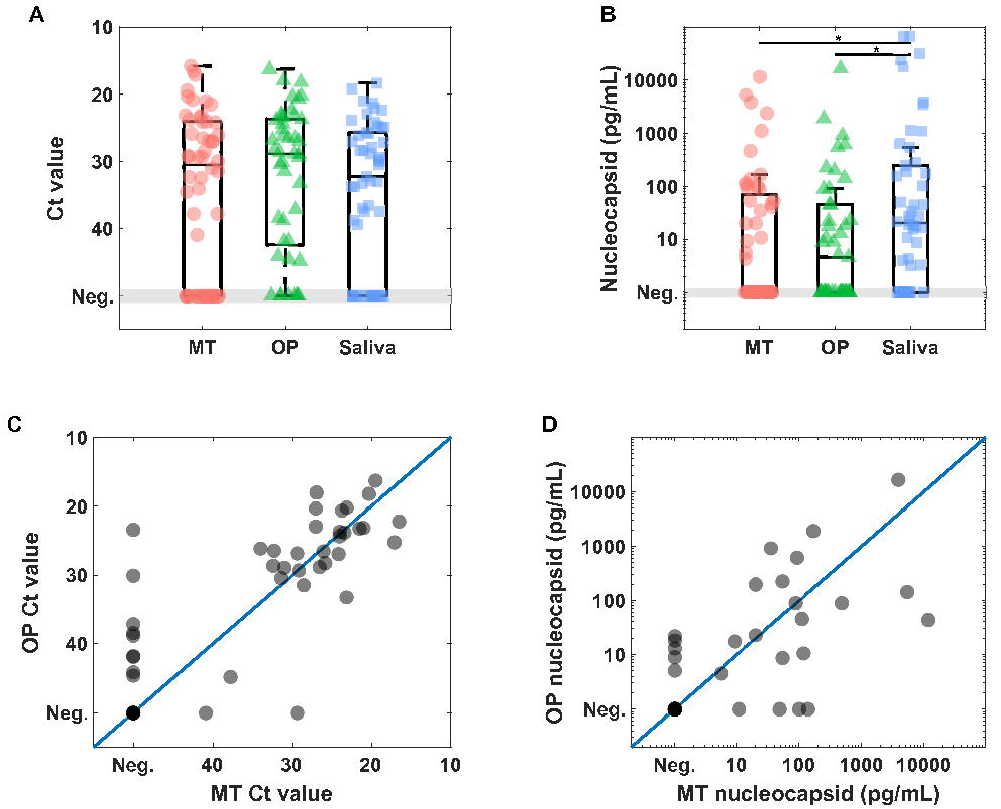
Time-independent comparison of (A) Ct values and (B) antigen levels for the three sample types. Significant differences in distribution of values based on rank-sum test were only observed for the comparisons of antigen concentration between MT and Saliva (p = 0.02) as well as OP and Saliva (p = 0.03). MT vs OP plots demonstrate distribution of (C) Ct values and (D) antigen concentration for paired specimens.

A Toeplitz covariance structure and an unstructured covariance matrix allowed the best fit model for the antigen concentrations and Ct values outcomes, respectively. Sample type was significantly associated with mean antigen concentrations (p = 0.04; **Table 2**), but this was not the case with Ct values (**Table 3**).

**Table 2.**
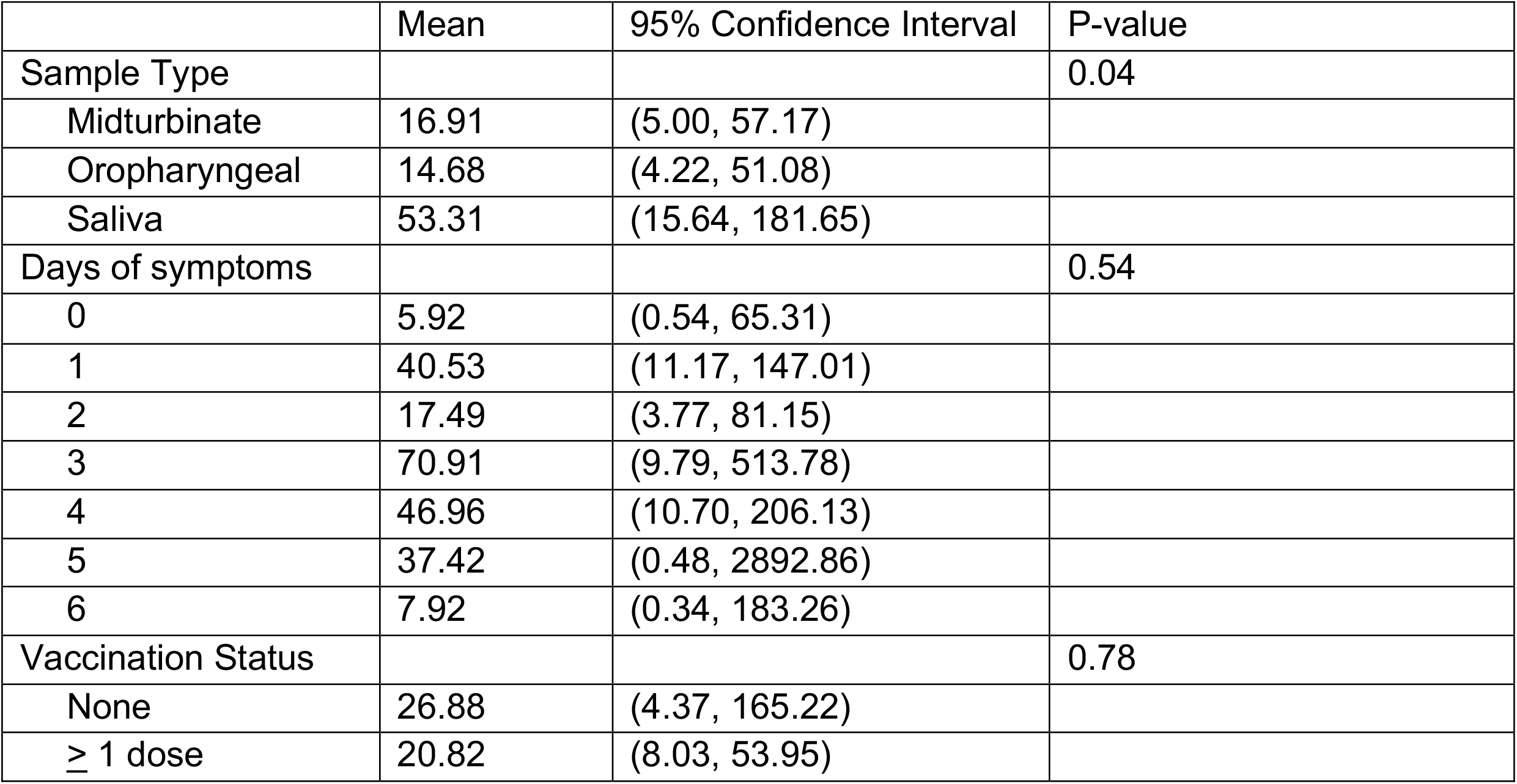
Impact of sample type and duration of symptoms on antigen concentrations using a mixed effect linear model (n=49)

**Table 3.**
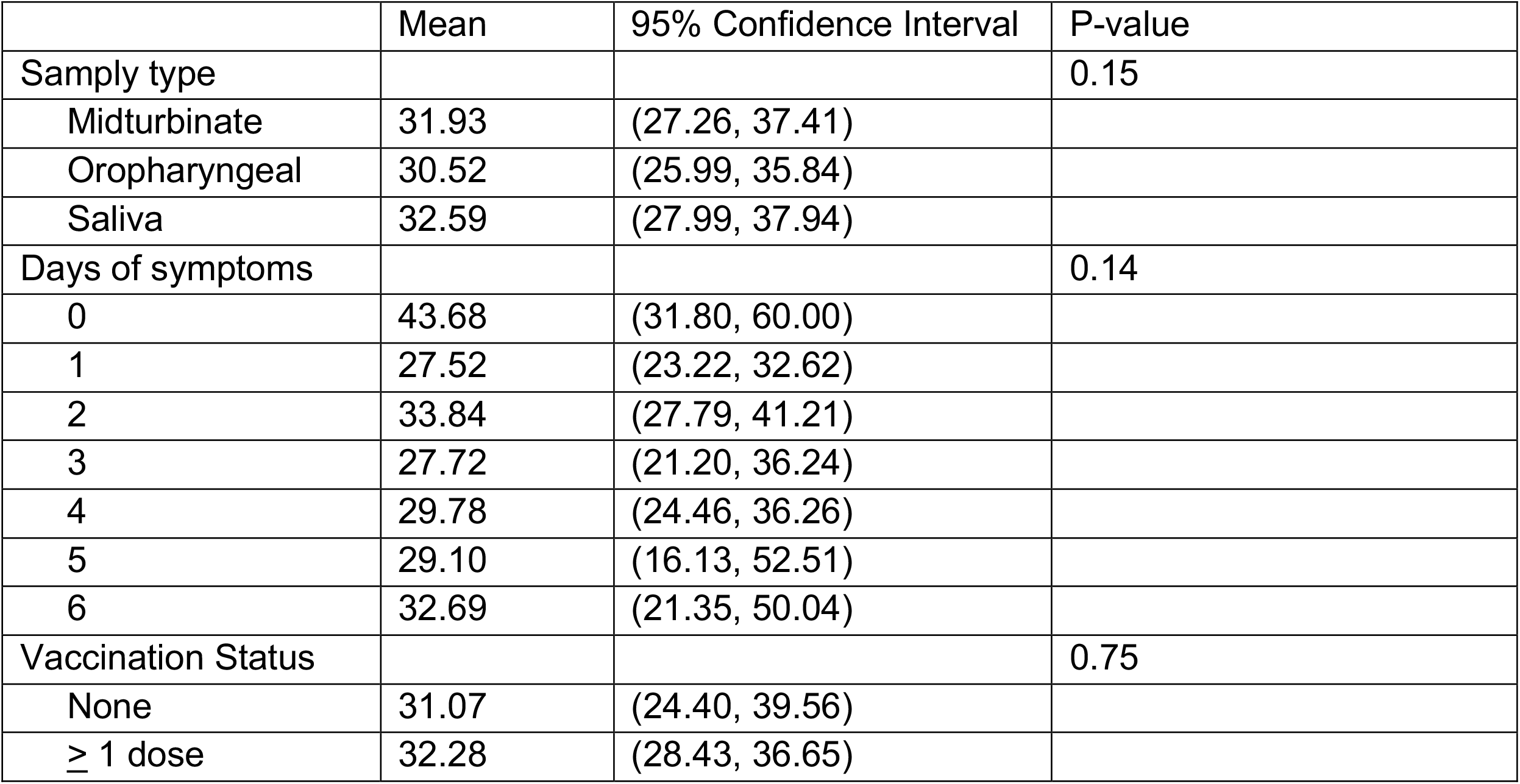
Impact of sample type and duration of symptoms on Ct values using a mixed effect linear model (n=49).

|  | Mean | 95% Confidence Interval | P-value |
| --- | --- | --- | --- |
| Sample type |  |  | 0.15 |
| Midturbinate | 31.93 | (27.26, 37.41) |  |
| Oropharyngeal | 30.52 | (25.99, 35.84) |  |
| Saliva | 32.59 | (27.99, 37.94) |  |
| Days of symptoms |  |  | 0.14 |
| 0 | 43.68 | (31.80, 60.00) |  |
| 1 | 27.52 | (23.22, 32.62) |  |
| 2 | 33.84 | (27.79, 41.21) |  |
| 3 | 27.72 | (21.20, 36.24) |  |
| 4 | 29.78 | (24.46, 36.26) |  |
| 5 | 29.10 | (16.13, 52.51) |  |
| 6 | 32.69 | (21.35, 50.04) |  |
| Vaccination Status |  |  | 0.75 |
| None | 31.07 | (24.40, 39.56) |  |
| $\geq 1$ dose | 32.28 | (28.43, 36.65) | |

Antigen concentrations and Ct values from three differing locations showed poor absolute agreement as determined by interclass correlation (ICC [95% CI] 0.12 [0.07–0.18] and 0.11 [0.07– 0.15], respectively). Ct values for OP and MT were directly compared (**Figure 3C-D**) as these sample types have identical collection methods. Ct value and antigen measurements both revealed a mix of samples with greater concentration in the OP sample (OP-predominant phenotype) and greater concentration in the MT sample (MT-predominant phenotype).

### Investigation of possible confounding factors

Ct value and antigen concentrations were examined separately based on duration of symptoms at time of sampling. Comparisons of Ct or antigen distribution did not differ significantly between sample types on any day of symptoms based on rank-sum test comparing OP and MT, OP and saliva, and MT and saliva (all p values > 0.05; Figure 4). The Toeplitz covariance structure and unstructured covariance matrix for antigen concentrations and Ct values, respectively, found no evidence that duration of symptoms nor vaccination status were significantly associated with antigen concentrations (p = 0.54 and 0.78, respectively; Table 2) or Ct value (p = 0.15 and 0.14, respectively; Table 3). Sensitivity analysis excluding negative Ct values that were imputed as 50 in the main analysis showed similar results (Table S2).

**Figure 4.**
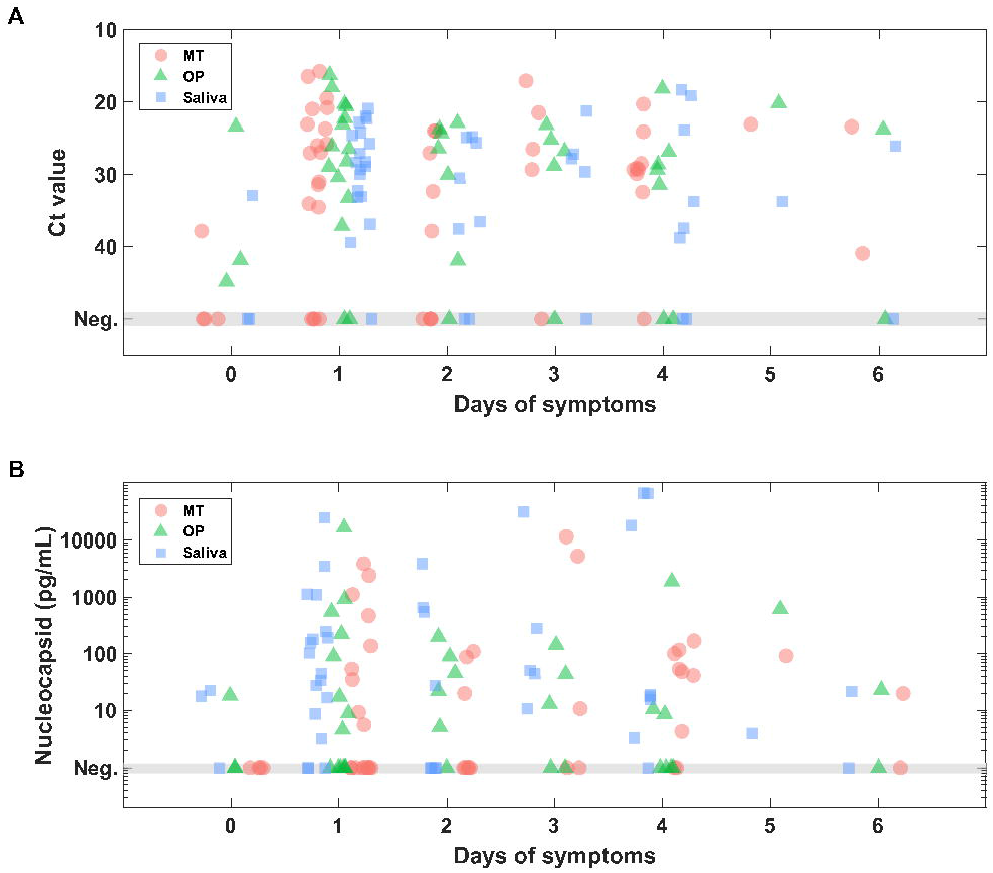
(A) Ct values and (B) nucleocapsid concentration sorted by duration of symptoms at time of sample collection.

## Discussion

Trends in self-testing for COVID-19 have raised interest in understanding typical viral loads of SARS-CoV-2 in specimens obtained from distinct anatomic sites within the upper respiratory tract. We collected specimens from a familial cohort of 3 individuals following exposure to SARS-CoV-2 and 121 symptomatic individuals presenting for testing, of which 54 had positive RT-PCR or antigen in at least one specimen.

Longitudinal samples from the familial cohort showed highest viral load in self-collected AN specimens compared to self-collected OP and saliva specimens for all three individuals. Plotting these measurements show that the curves rarely cross, suggesting that there were similar viral kinetics in all three sample types for these individuals and that the AN consistently remained the site of highest viral load. As such, this cohort appeared to maintain a consistent nasal-predominant phenotype among samples collected by non-healthcare trained participants.

Although all three members of the familial cohort exhibited a consistent nasal predominance in viral load, the cross-sectional cohort demonstrated a mix of MT-predominant and OP-predominant phenotypes and there was not a sample type with consistently higher viral RNA and/or nucleocapsid antigen across the entire cohort. Our data did not provide evidence that one sample type was more reliable for clinical testing owing to significant overlap in diagnostic performance of all three sample types. Throughout the range of antigen concentrations observed and including the range at which common OTC RDTs perform, there appears to be no advantage to using one sample type relative to the other (**Figure 2B**).

Limitations of our data include the use of self-collected swabs in the familial cohort, which may not have been collected as reliably as those collected by a healthcare provider in the cross-sectional cohort. RT-PCR assays were performed with a common assay (Cepheid GeneXpert) for MT and OP samples while saliva was assayed based on the SalivaDirect protocol. Neither assay has EUA approval as a quantitative test and this may limit conclusions, particularly from comparison of Ct values between saliva and the other specimens.

Additionally, the quantity of cellular material and mucous may be inherently different between sample types with different collection methodologies, particularly with saliva collected as a pooled drool specimen compared to MT and OP samples obtained with collection swabs eluted in buffer. Despite this, the pragmatic question of viral yield for diagnostic samples can be investigated in this manner particularly because a consistent sample volume and a common assay was used for nucleocapsid measurements across all sample types. Thus, our data appear to suggest that higher antigen levels may be found in saliva specimens, but calculation of PPA failed to show benefit in diagnostic yield. Conclusions as to the utility of saliva for antigen testing may be limited by missing specimens from some participants and a small sample size.

The observation of OP- and MT-predominant phenotypes is a key finding in our data in addition to the absence of an effect of days of symptoms at the time of sampling. Isolated reports of OP-positive, nares-negative tests early during symptomatic COVID-19 and practices of combined oropharyngeal and nasal sampling are among the factors motivating this investigation. Testing of 16 individuals in our study (8 positive by another method, 8 negative by all alternative tests) with separate AN and OP specimens on a common OTC POC test (Quickvue OTC home test) showed 3/8 true positives from the AN, 5/8 false negatives from the AN, 8/8 false negatives from the OP and no false positives for any sample type. One hypothesis compatible with our data is that many individuals with COVID-19 in late 2021 and early 2022 manifest either OP- or MT-predominant viral loads in the upper respiratory tract. Ongoing examination of this question including larger sample sizes is warranted, particularly if new variants of concern were to emerge.

In total, our data do not support a preferred sample type for SARS-CoV-2 detection during the omicron era but suggest heterogenous distribution of viral loads in MT, OP, and saliva sample types collected from symptomatic persons with COVID-19 during the omicron era. Further rigorous study should investigate the benefit and safety of combined sampling methods.

## Conclusions

Effective testing strategies are an essential tool to limit the spread of highly transmissible disease. The emergence of the SARS-CoV-2 omicron variant created uncertainty in appropriate testing measures, leading to variability in testing techniques that risk the efficacy of validated tests and sample collection methods. Our data suggest that SARS-CoV-2 nucleocapsid and RNA exhibit similar kinetics and diagnostic yield in three upper respiratory sample types across the duration of symptomatic disease. Collection of OP or combined nasal and OP samples does not appear to increase sensitivity versus validated nasal sampling for rapid detection of viral antigen. We therefore advise against changing recommended testing practices until additional information is discovered.

## Data Availability

All data produced in the present study are available upon reasonable request to the authors

## Notes

This work was supported by the National Institute of Biomedical Imaging and Bioengineering at the National Institutes of Health under award Numbers U54 EB027690-03S1 and U54 EB027690-03S2 and the National Center for Advancing Translational Sciences of the National Institutes of Health under Award Number UL1TR002378. The content is solely the responsibility of the authors and does not necessarily represent the official views of the National Institutes of Health.

### Competing Interest Statement

The authors have declared no competing interest.

### Author Declarations

IRB of Emory University gave ethical approval for this work

## References

1. Marty, F. M., Chen, K. & Verrill, K. A. How to Obtain a Nasopharyngeal Swab Specimen. N. Engl. J. Med. 382, e76 (2020).

2. Nikolai, O. et al. Anterior nasal versus nasal mid-turbinate sampling for a SARS-CoV-2 antigen-detecting rapid test: does localisation or professional collection matter? Infect. Dis. 53, 947–952 (2021).

3. Savela, E. S. et al. Quantitative SARS-CoV-2 viral-load curves in paired saliva and nasal swabs inform appropriate respiratory sampling site and analytical test sensitivity required for earliest viral detection. J. Clin. Microbiol. (2021) doi:10.1128/JCM.01785-21.

4. Wong, S. C. Y. et al. Posterior Oropharyngeal Saliva for the Detection of Severe Acute Respiratory Syndrome Coronavirus 2 (SARS-CoV-2). Clin. Infect. Dis. 71, 2939–2946 (2020).

5. Echavarria, M. et al. Self-collected saliva for SARS-CoV-2 detection: A prospective study in the emergency room. J. Med. Virol. 93, 3268–3272 (2021).

6. Saegerman, C. et al. Repetitive saliva-based mass screening as a tool for controlling SARS-CoV-2 transmission in nursing homes. Transbound. Emerg. Dis. n/a,.

7. Ricci, S. et al. How home anterior self-collected nasal swab simplifies SARS-CoV-2 testing: new surveillance horizons in public health and beyond. Virol. J. 18, 59 (2021).

8. Valentine-Graves, M. et al. At-home self-collection of saliva, oropharyngeal swabs and dried blood spots for SARS-CoV-2 diagnosis and serology: Post-collection acceptability of specimen collection process and patient confidence in specimens. PLOS ONE 15, e0236775 (2020).

9. Health, C. for D. and R. In Vitro Diagnostics EUAs - Antigen Diagnostic Tests for SARS-CoV-2. FDA (2022).

10. CDC. COVID-19 and Your Health. Centers for Disease Control and Prevention https://www.cdc.gov/coronavirus/2019-ncov/testing/self-testing.html (2020).

11. How to do a rapid lateral flow test for coronavirus (COVID-19). nhs.uk https://www.nhs.uk/conditions/coronavirus-covid-19/testing/how-to-do-a-test-at-home-or-at-a-test-site/how-to-do-a-rapid-lateral-flow-test/ (2021).

12. Tsang, N. N. Y. et al. Diagnostic performance of different sampling approaches for SARS-CoV-2 RT-PCR testing: a systematic review and meta-analysis. Lancet Infect. Dis. 21, 1233–1245 (2021).

13. Lee, R. A., Herigon, J. C., Benedetti, A., Pollock, N. R. & Denkinger, C. M. Performance of Saliva, Oropharyngeal Swabs, and Nasal Swabs for SARS-CoV-2 Molecular Detection: a Systematic Review and Meta-analysis. J. Clin. Microbiol. (2021) doi:10.1128/JCM.02881-20.

14. Zhou, Y. & O’Leary, T. J. Relative sensitivity of anterior nares and nasopharyngeal swabs for initial detection of SARS-CoV-2 in ambulatory patients: Rapid review and meta-analysis. PLOS ONE 16, e0254559 (2021).

15. Nsoga, M. T. N. et al. Diagnostic accuracy of Panbio rapid antigen tests on oropharyngeal swabs for detection of SARS-CoV-2. PLOS ONE 16, e0253321 (2021).

16. Agulló, V. et al. Evaluation of the rapid antigen test Panbio COVID-19 in saliva and nasal swabs in a population-based point-of-care study. J. Infect. 82, 186–230 (2021).

