## Supplement for "Where is Omicron? Comparison of SARS-CoV-2 RT-PCR and Antigen Test Sensitivity at Commonly Sampled Anatomic Sites Over the Course of Disease"

Supplementary Information

| **OTC test** | **LOD (pg/mL)** |
| --- | --- |
| QuickVue At-Home OTC COVID-19 Test | 2,749 |
| BinaxNOW COVID-19 Antigen Self Test | 1,441 |
| BD Veritor At-Home COVID-19 Test | 569 |
| MaximBio ClearDetect COVID-19 Antigen Home Test | 150 |

**Table S1.** Expected cutoffs of common OTC rapid diagnostic tests presented in Figure 2B. These values were determined by rangefinding studies where LOD (limit of detection) was defined as the lowest tested antigen concentration at which 100% of 5 replicates produced a positive result.

|  | **Mean** | **95% Confidence Interval** | **P-value** |
| --- | --- | --- | --- |
| Sample type |  |  | 0.12 |
| Midturbinate | 27.94 | (24.86, 31.39) |  |
| Oropharyngeal | 27.72 | (24.71, 31.09) |  |
| Saliva | 30.16 | (26.87, 33.86) |  |
| Duration of symptoms |  |  | 0.21 |
| 0 | 37.27 | (28.25, 49.16) |  |
| 1 | 26.19 | (23.29, 29.45) |  |
| 2 | 29.41 | (25.41, 34.04) |  |
| 3 | 24.72 | (20.34, 30.04) |  |
| 4 | 27.17 | (23.59, 31.29) |  |
| 5 | 27.52 | (18.61, 40.70) |  |
| 6 | 29.39 | (21.43, 40.30) |  |
| Vaccination Status |  |  | 0.91 |
| Received at least 1 dose | 28.46 | (25.98, 31.17) |  |
| Has never received a COVID-19 vaccine dose | 28.71 | (24.26, 33.98) |  |

**Table S2.** Sensitivity Analysis: Impact of sample type and duration of symptoms on Ct values using a mixed effect linear model excluding observations with a negative Ct value (N=46).
